# Heterogeneity in elevated glucose and A1C as predictors of the prediabetes to diabetes transition: Framingham Heart Study, Multi-Ethnic Study on Atherosclerosis, Jackson Heart Study, and Atherosclerosis Risk In Communities

**DOI:** 10.1101/2024.03.16.24304398

**Authors:** Chirag J. Patel, John PA Ioannidis, Edward W. Gregg, Ramachandran S. Vasan, Arjun K. Manrai

**Author notes:** Correspondence to: Chirag J Patel.

## Abstract

**Introduction:** There are a number of glycemic definitions for prediabetes; however, the heterogeneity in diabetes transition rates from prediabetes across different glycemic definitions in major US cohorts has been unexplored. We estimate the variability in risk and relative risk of adiposity based on diagnostic criteria like fasting glucose and hemoglobin A1C% (HA1C%).

**Research Design and Methods:** We estimated transition rate from prediabetes, as defined by fasting glucose between 100-125 and/or 110-125 mg/dL, and HA1C% between 5.7-6.5% in participant data from the Framingham Heart Study, Multi-Ethnic Study on Atherosclerosis, Atherosclerosis Risk in Communities, and the Jackson Heart Study. We estimated the heterogeneity and prediction interval across cohorts, stratifying by age, sex, and body mass index. For individuals who were prediabetic, we estimated the relative risk for obesity, blood pressure, education, age, and sex for diabetes.

**Results:** There is substantial heterogeneity in diabetes transition rates across cohorts and prediabetes definitions with large prediction intervals. We observed the highest range of rates in individuals with fasting glucose of 110-125 mg/dL ranging from 2-18 per 100 person-years. Across different cohorts, the association obesity or hypertension in the progression to diabetes was consistent, yet it varied in magnitude. We provide a database of transition rates across subgroups and cohorts for comparison in future studies.

**Conclusion:** The absolute transition rate from prediabetes to diabetes significantly depends on cohort and prediabetes definitions.

**Twitter Summary:** New study finds variable diabetes risk in prediabetes across US cohorts. Results highlight obesity, Black race, and hypertension as key factors, emphasizing the need for precision in diabetes care. #DiabetesResearch

---

The prevalence of prediabetes is genuinely heterogeneous across the United States (1,2). Estimates of diabetes risk depend strongly on the chosen analytes (e.g., fasting glucose, 2 hour glucose, HbA_1C_) and cut-points, with multiple definitions currently in use from the American Diabetes Association (ADA) and World Health Organization (WHO) (3). The ADA, in their newest standards of care, have recommended screening for prediabetes for any of these analytes. The ADA also recommends cardiovascular preventive measures (e.g., lifestyle change and weight loss (4)) who are prediabetics, again, under any of the definitions of prediabetes (5).

While prediabetes presents as a modifiable risk for diabetes, the definition of prediabetes relates to cohort-level sources of variation and predicts the transition from prediabetes to overt diabetes has not been well characterized. One study of US-based participants of a claims database estimated that the annual incidence rate (the number of individuals with diabetes diagnoses per person-time)), ranged between 2.5-8.2 per 100 person-years for A1C-based definitions (6). Another study, in participants of the final exams of the Atherosclerosis Risk in Communities (ARIC) cohort recruited from communities from Minnesota, Maryland, Mississippi, and North Carolina, found a range of 1.4-1.9 and 1.6-2.2 per 100 person-years for FBG- and A1C-based definitions, respectively, in the >65 year old population (7).

Here, we study the range and heterogeneity in the absolute incidence and the relative risk for diabetes across competing definitions of prediabetes in participants from four large US cohorts, including the Framingham Heart Study (FHS) Offspring (“Offspring”) study (8), FHS Generation three (“Generation 3”), Multi-Ethnic Study on Atherosclerosis (“MESA”) (9), Atherosclerosis Risk in Communities (“ARIC”) (10) and the Jackson Heart Study (“JHS”) (11). These cohorts have recruited from four race/ethnic groups, and they vary in the age groups that they cover, the calendar period of recruitment and follow-up (1975-2013), and the distributions of clinical risk phenotypes (e.g., body mass index and blood pressure).

We analyze the range of the risk of transition to diabetes based on cohort, definitions of prediabetes (e.g., glucose levels based FBG or A1C), and baseline age at inclusion criteria. We also assess the role of time of cohort recruitment. We also estimate the role of competing and complementary individual-level clinical risk factors that are also used in screening, such as body mass index.

## Research Design and Methods

We have followed the STrengthening the Reporting of OBservational studies in Epidemiology (STROBE) checklist in reporting of the observational study design findings.

### Study cohorts and design

We performed a cohort-level analysis of five community-based longitudinal cohorts: the Framingham Heart Study (FHS) Offspring (“Offspring”) study, FHS Generation three (“Generation 3”), Multi-Ethnic Study on Atherosclerosis (“MESA”), Atherosclerosis Risk in Communities (“ARIC”) and the Jackson Heart Study (“JHS”). We downloaded cohort data from NHLBI BioLINCC as of December 2020. The FHS Offspring cohort consisted of participants sampled from Framingham, Massachusetts in 6 serial exams occurring during 1983-2014 (Exam 3: 1983-1987; Exam 4: 1987-1991; Exam 5: 1991-1995; Exam 6: 1995-1998; Exam 8: 2005-2008; Exam 9: 2011-2014). FHS Generation 3 consisted of 2 serial exams from 2002-2011 (Exam 1: 2002-2005; Exam 2: 2008-2011; Exam 3: 2016-2019). MESA consisted of 5 exams during 2000-2011, sampling participants from 6 communities from New York, Baltimore, Chicago, Los Angeles, Twin Cities, and Winston-Salem (Exam 1: 2000-2002; Exam 2: 2002-2004; Exam 3: 2004-2005; Exam 4: 2005-2007; Exam 5: 2010-2011). ARIC consisted of 5 exams during 1987-2012 from 4 communities, including individuals from Washington County, Maryland; Forsyth County, North Carolina; Jackson, Mississippi; and Minneapolis, Minnesota (Exam 1: 1987-1989; Exam 2: 1990-1992; Exam 3: 1993-1995; Exam 4: 1996-1998; Exam 5: 2009-2012). The JHS consisted of 3 serial exams from 2000-2013 (Exam 1: 2000-2004; Exam 2: 2005-2008; Exam 3: 2009-2013). Some participants of JHS also are part of MESA. We removed participants of both JHS and MESA from the MESA cohort for all subsequent analyses, to ensure non-overlap in participants. Our inclusion criteria included all participants examined in at least two exams aged 18 or older.

### Definitions of incident diabetes diagnosis

In all cohorts, our main outcome of interest was incident diabetes. In the FHS Offspring and Generation 3 studies, diabetes was classified by either having fasting blood glucose greater than or equal to 126 mg/dL or being under treatment for diabetes. In MESA, diabetes was diagnosed as having fasting blood glucose greater than or equal to 126 mg/dL, or being under treatment for diabetes. In ARIC, diabetes was diagnosed as fasting blood glucose greater than or equal to 126 mg/dL or being under treatment for diabetes. In JHS, diabetes was classified as fasting blood glucose greater than 126 mg/dL or a 2-hour blood glucose level in response to an oral glucose tolerance test (OGTT) of greater than 200 mg/dL, or being under treatment for diabetes.

We also sought to compare incident total diabetes to incident high fasting glucose or HA1C% (regardless of treatment). We determined high fasting glucose as those incident cases that exceed 126 mg/dL and/or HA1C greater than 6.5%; incident high fasting glucose; incident high A1C%; and incident both high A1C and fasting glucose.

### Prediabetes, obesity, and hypertension definitions and inclusion criteria

We considered the American Diabetes Association (ADA) range for prediabetes (100-125 mg/dL) and the World Health Organization (WHO) range for prediabetes (110-125 mg/dL) for Framingham Generation 2, Framingham Generation 3, MESA, ARIC, and JHS cohorts. We considered glycated hemoglobin percent (A1C) between 5.7% and 6.4% for subsets in the Framingham Generation 2, MESA, ARIC, and JHS cohorts, where this information was available. We also considered individuals who only fell into one (e.g., only fasting 100-125 mg/dL, 110-125 mg/dL, or HA1C 5.7-6.4%) or any two (e.g., 110-125 mg/dL AND 5.7-6.4). We defined clinical overweight and obesity categories using body mass index, specifically, obesity (>= 30kg/m2), overweight (25-30 kg/m2), and non-obesity/overweight (0-25 kg/m2). We also defined hypertension as having either systolic or diastolic blood pressure greater than 140 mm Hg or 90 mm Hg respectively. We defined high blood pressure as having either systolic or diastolic blood pressure greater than 130 or 80 mm Hg.

### Cohort-specific prediabetes incidence rates for diabetes

For each prediabetes definition (n=3) and age group (n=3, [year ranges 18-44, 45-64, 64-100]) we estimated the age- and sex-adjusted incidence rate or risk for diabetes for each cohort separately. To estimate age- and sex-adjusted rates, we used a multivariable Poisson regression model to calculate predicted diabetes counts (with a logarithm of follow-up time offset term). Within cohort subgroups included body mass index (obesity: >=30 kg/m2, overweight: 25-30 kg/m2, and non-obesity/overweight: 0-25 kg/m2), race (MESA: White, Black, Hispanic, Chinese; ARIC: White/Black; JHS: Black; FHS: White), and reported sex (male/female).

To estimate the incidence rate, we weighted the predicted count by the frequency of the type of individual in the population (4) and bootstrapped the cohorts (100 samples) to estimate the standard error of the rate. All incidence rates are reported as annual percentage, or number of cases per 100 person-years at risk.

We estimated the overall and heterogeneity of the incidence rates using a random-effects model (restricted maximum likelihood estimation) across cohorts. We also estimated a two-sided 95% prediction interval (PI) for the incidence rate across all cohorts (12). The usual overall incidence rate estimated using a random effects model is the average effect across studies. The prediction interval, on the other hand, is the predicted range of the incidence rate of prediabetes to diabetes for a new cohort.

### Data-driven thresholds for fasting glucose and A1C%

Aside from a priori defined thresholds for prediabetes, we also estimated thresholds that maximized accuracy of prediction of incident diabetes, incident glucose greater than 126 mg/dL, and incident A1C% greater than 6.5% as a function of baseline fasting glucose and A1C%. We generated a Receiver Operating Characteristic (ROC) curve (sensitivity versus specificity) and selected the threshold along the curve that maximizes accuracy (fraction of correctly classified incident cases). We drew 100 bootstrap samples to estimate the standard error of the thresholds that maximized accuracy.

### Modeling prediabetes to diabetes relative risk and assessing inter-cohort heterogeneity across clinical and sociodemographic variables

To assess the role of clinical (e.g., body mass index, blood pressure), behavioral (smoking), and sociodemographic factors (age, sex, race, and education) in the relative risk for diabetes from prediabetes, we performed cohort-specific multivariable regression analyses using a Cox proportional hazards model. Specifically, for each cohort, we chose entry criteria for individuals who had FBG (a) lower than 100 mg/dL, (b) between 100-125 mg/dL, (c) lower than 110 mg/dL, (d) between 110-125mg/dL, or (e) who had A1C between 5.7% and 6.4%. We estimated the cross-cohort hazard ratios for each of these prediabetes definitions using a random-effects model (restricted maximum likelihood estimation).

## Results

We refer to individuals with prediabetes as having high fasting glucose (two groups, including 100-125 mg/dL [ADA] or 110-125 mg/dL [WHO]) or glycated hemoglobin A1C percent (5.7-6.4% [A1C]). Each cohort had a different baseline case-mix that reflected differences in the prevalence of high fasting glucose, A1C% and cardiometabolic risk factors, such as obesity and to a smaller degree, hypertension (Table S1-3)

15% of participants of FHS 2 had ADA prediabetes (and 3.4% had WHO prediabetes) whereas the FHS Generation 3 (children of the FHS 2) had an ADA prediabetes prevalence of 20%. ARIC, whose participants were approximately 10 years older versus both FHS cohorts, had a 44% prevalence of ADA prediabetes (13% with WHO prediabetes). Participants of JHS also had a lower prevalence of ADA and WHO prediabetes (10 and 2.2%, respectively). In contrast, the prevalence of A1C defined prediabetes was almost 4 times greater for JHS participants, 36%, in comparison to ADA FBG-defined prediabetes. We also observed greater prevalence of A1C defined diabetes in MESA participants. JHS participants had a much higher prevalence of obesity relative to FHS Generation 2, FHS Generation 3, and ARIC cohorts, 75% versus 30-38% (Table S2). MESA had the next largest prevalence of obesity (42%) vis-à-vis prevalence of ADA prediabetes.

### Baseline fasting glucose and A1C across the cohort, BMI, race categories

There were differences in the spectrum of fasting glucose across cohorts and across BMI categories within each cohort (Figure 1), and also across subgroups of race, age and sex (Figure S1) measured at baseline. The median FBG levels for individuals in ARIC were 100, whereas the median FBG in the other cohorts were 10-15 mg/dL lower (Figure 1A) and “outside” the range for FBG-defined prediabetes. On the other hand, participants in all cohorts had A1C well below the range A1C-defined prediabetes (Figure 1C). 40% of the obese population had fasting glucose levels in the 100-125 mg/dL range(Figure 1B), whereas approximately 25% of the obese population had A1C% in the prediabetic range (Figure 1D). Black and Hispanic individuals had lower median glucose levels than white and Chinese (Figure S1). The older and younger populations had higher FBG than their mid age (44-64 years of age) counterparts (Figure S1A). Men had higher median fasting glucose levels (Figure S1 C, F).

**Figure 1.**
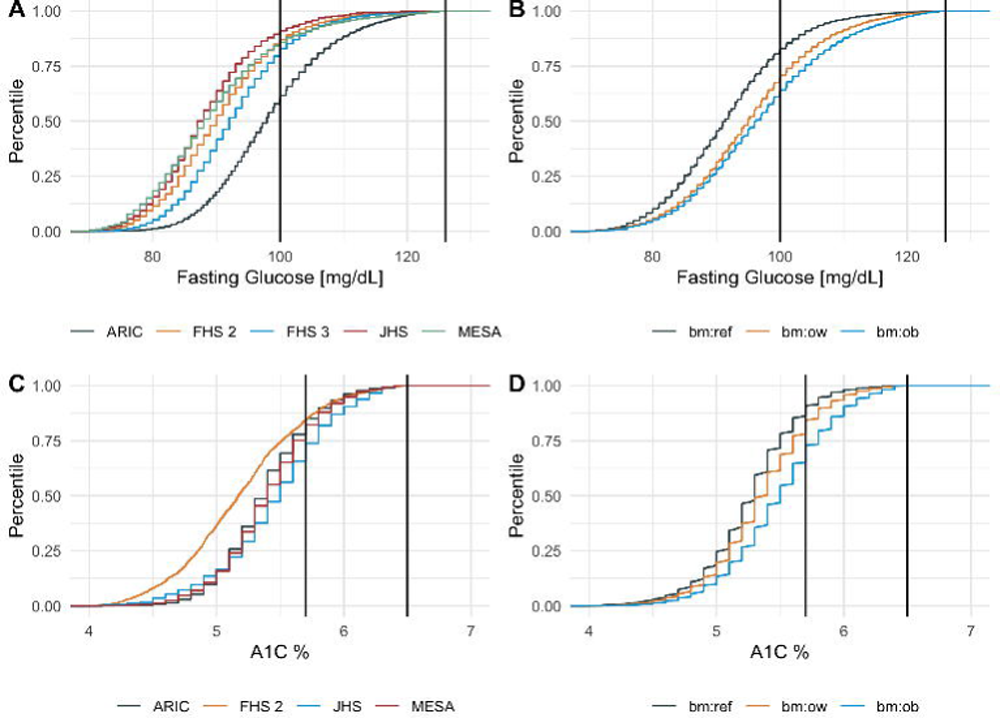
Distributions of baseline fasting glucose and A1C by A.) Cohort (FHS Generation 2, FHS Generation 3, ARIC, JHS, MESA), B.) BMI cutoffs (bm:ref denotes 0-25 kg/m2, bm:ow denotes 25-30 kg/m2; bm:ob denotes greater than 30 kg/m2).

### Incidence rates within and across cohorts for incident diabetes

We estimated the incidence rates for (1) diabetes (higher fasting/non-fasting glucose and/or glucose lowering drug) and (2) higher fasting glucose (greater than or equal to 126 mg/dL) and (3) higher A1C% (greater than or equal to 6.5%).

Incidence rates for all three incident outcomes were dependent on cohort and prediabetes definition. First, we discuss incident diabetes. The cross-cohort meta-analytic estimate (adjusted by age and sex and averaged using a random-effects meta-analysis) rate for transition to diabetes for FBG 100-125 mg/dL, 110-125 mg/dL, and A1C 5.7-6.4% were 4.2 per 100 person-years, or 4.2% (95% Confidence Interval [CI]: 1.7-6.8; 95% Prediction Interval [PI]: −4.5-13), 8.3% (95% CI: 4.0-12.6; 95% PI: −6.5-23%), 2.8% (95% CI: 1.2-4.4, 95% PI: −3.0-8.6) cases per 100 person-years, respectively (Figure 2). This contrasts with incidence rates of 0.6% (95% CI: 0.01-1.2; 95% PI: 1.4-2.6), 1.0% (95% CI: 0.2-1.7; 95% PI: −1.6-3.5), and 0.6% (95% CI: 0.45-0.70; 95% PI: 0.2-1%) for the individuals with FBG less than 100, FBG less than 110, and A1C% less than 5.7% respectively. However, the range of the incidence rates across cohorts was large (Figure 2). All the meta-analytic estimates were the I^2^ for all prediabetes thresholds were greater than 93% (Figure 2) and for normoglycemic thresholds were greater than 76%. The prediction intervals ranged from below 1 up to 23% (Figure 2, red dotted line), indicating that we would expect to see any new transition rate from a new cohort sample to be in this range.

**Figure 2.**
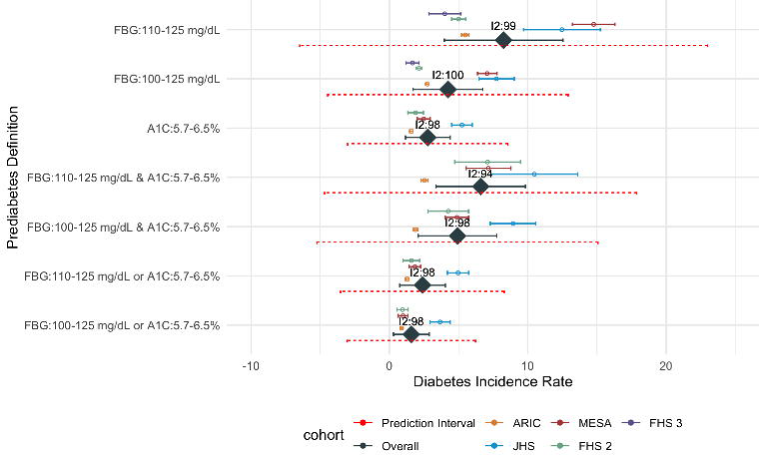
Cohort-specific and meta-analytic (cross-cohort) incidence/transition rates (per 100 person-years) for diabetes for different definitions of prediabetes, including non-elevated and elevated FBG and HA1C%. Colors for open symbol points denote cohort, JHS: Jackson Heart Study; FHS 2: Framingham Generation 2 (Offspring); FHS 3: Framingham Generation 3; MESA: Multi-Ethnic Study on Atherosclerosis; ARIC: Atherosclerosis Risk in Communities. Overall: Overall random-effect meta-analytic estimate across cohorts (in solid purple). The 95% prediction intervals are seen in the dotted red line.

The between-cohort range of the incidence rates were large. For example, or FBG 110-125 mg/dL was 4% (95% CI: 2.9-5.2) for FHS Generation 3, 5% (95% CI: 4.5-5.5) for FHS Generation 2, 5.5% (95%CI: 5.2-5.8) for ARIC, 12.5% (95% CI: 9.7-15.3) for JHS, and 14.8% (95% CI: 13.2-16.3) for MESA (Figure 2, open points). In contrast, the range of the incidence rate for A1C 5.7-6.4% was 1.6% (95% CI: 1.5-1.7) for ARIC and 5.2% (95% CI: 4.4-6.0 for JHS, markedly lower.

We also estimated incidence rates for individuals with different combinations of glycemic definitions, such as having both a higher fasting glucose and A1C% versus just one. The heterogeneity of the incidence rate did not diminish for individuals who had both elevated fasting and HA1C levels. For example, the cross-cohort range for the incidence rates for individuals with both fasting glucose between 110-125 mg/dL and A1C% between 5.7 and 6.4% was 2.5 per 100 person years (2.5%, ARIC), 7.1% (FHS 2), 7.2% (MESA), and 11% (JHS). The overall meta-analytic transition rate for this group was 6.6 (95% CI: 3.3-9.9; 95% PI: −4.7-17.9; I^2^ = 94%). For individuals with both lower fasting glucose and lower A1C% was 0.5% (95% CI: 0.3-0.6; 95% PI: −0.2-1.1; I^2^ of 91%)

We also conducted sensitivity analysis to assess whether diabetes diagnosis definitions across cohorts may contribute to heterogeneity. Cross-cohort heterogeneity for FBG-defined prediabetes persisted when considering incident FBG greater than 126 mg/dL or A1C% greater than 6.5% (Figure S2). For example, for a prediabetes defined by 110-126 mg/dL, the incidence rates were 2.7% (FHS 3), 4% (FHS 2), 4% (JHS), 5% (ARIC), and 10.5% (MESA). The overall meta-analytic incidence for FBG greater than 126 mg/dL was 5.1% (I^2^: 99%).

### Incidence rates for age, sex, body mass index, and race subgroups

We identified differences in rates across risk subgroups, including age (Figure S3A, Figure S4A), body mass index categories (Figure S3B, Figure S4B), sex (Figure S5), and race (Figure S6). The distributions of incidence rate estimates for ARIC, and FHS cohorts overlap but are generally lower than MESA and JHS (Figure 3). We release all estimated subgroup incidence rates and their standard errors as a supplementary table (Table S4). The Black subgroup was associated with the highest rate of diabetes, especially for individuals with FBG-defined prediabetes from the MESA and FHS cohorts (Figure S6). We provide a table of incidence rates across all subgroups and cohorts in Table S4 and a summary of the findings in the Supplementary Document.

**Figure 3.**
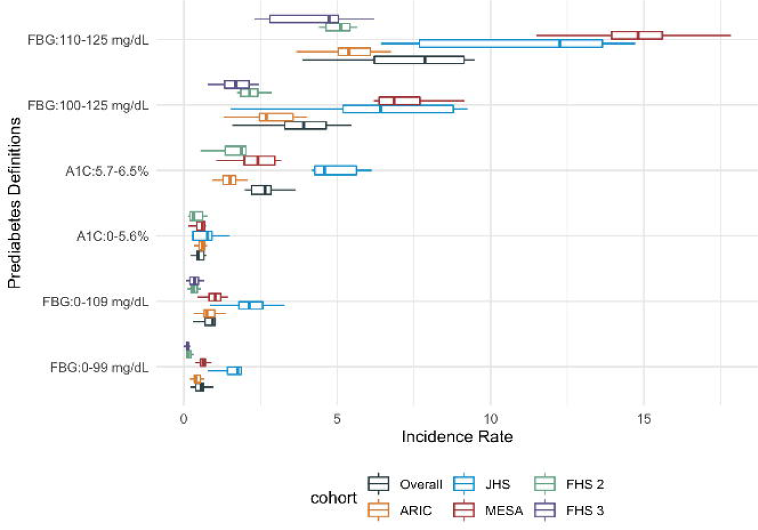
Distribution of subgroup incidence rates (per 100 person years) for diabetes by prediabetes definitions and cohort across different age, sex, and BMI groups.

### Data-driven thresholds

We estimated the “optimal” threshold for fasting glucose or HA1C% that maximized the accuracy of prediction of diabetes for each cohort (Table S5). We estimated the highest accuracy of for MESA (accuracy of 0.88 for FBG threshold of 103 mg/dL and 0.83 for a A1C% threshold of 5.8, We observed the lowest accuracy for JHS (accuracy of 0.71 for FBG threshold of 95, and accuracy of 0.73 for A1C threshold of 5.8. The thresholds that maximized accuracy exhibited heterogeneity of 65% for A1C (range of 5.5 to 5.7) and 91% for fasting glucose (range of 95 to 104 mg/dL). Predicted data driven thresholds were close to the fixed ADA thresholds.

### Within and between cohort heterogeneity of relative risk for diabetes

We estimated the within-cohort hazard ratio [HR] (or relative risk) for diabetes of sex, race/ethnicity (for ARIC and MESA), age, education, blood pressure, body mass index, and current smoking status using a Cox proportional hazards model with time-dependent covariates (Figure S7). Figure S8 describes the overall or meta-analytic estimates. We deemed an FDR level of 0.05 (p-value of 0.01, Figure S7-8) as significant, and those factors and HR are displayed in Figure 4.

**Figure 4.**
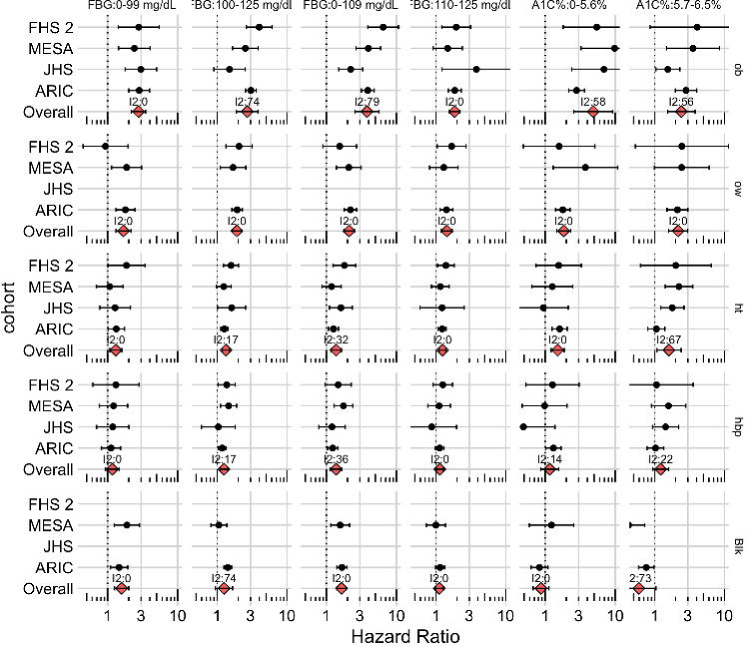
Multivariate relative risks for individuals with prediabetes and normoglycemia. Hazard ratio vs. p-value by cohort for individuals with prediabetes (FBG 100-125 mg/dL, 110-125 mg/dL, A1C 5.7-6.5%) or normal glycemia(FBG 0-99 mg/dL, 0-109 mg/dL or A1C% 0-5.7). I^2^ are labeled with the Overall estimate (diamond). Variables are labeled: Ob: BMI greater than 30 kg/m2 (ref: BMI between 0-25 kg/m2); ow: BMI 25-30 kg/m2 (ref: BMI between 0-25 kg/m2); ht: hypertension; hbp: high blood pressure (ref: normal blood pressure);; Blk: Black race (ref: vs. White).

We found obesity and overweight to be consistently highlighted for individuals regardless of FBG/A1C regardless of prediabetes status (Figure 4). First, we describe hazard ratios (HRs) found in individuals with prediabetes (e.g., FBG 100-125 mg/dL or A1C% 5.7-6.4). The range of the obesity HR (vs. non-obese) was 1.5 (MESA) to 4.0 (FHS Offspring), with the highest HR observed in the FHS Generation 2 cohort (adjusted HR: 4.0, 95% CI: 2.6, 6.1) in individuals with FBG 100-125 mg/dL. Second, of non-prediabetic or normoglycemic individuals, the HR ranged from 2.2 to 9.8, with the highest HR (9.8, 95% CI: 3.3, 29.6) found in MESA participants with obesity and lower A1C% (<5.7%) (Figure 4).

When combining HR across cohorts in a meta-analysis (Figure 4 [“Overall”], Figure S8), the role of factors such as hypertension, smoking, and overweight became more prominent in both prediabetic (Figure 4, Figure S8A). For example, across all cohorts, individuals with A1C- or FBG-defined prediabetes with hypertension have a 40-60% increased risk for diabetes (max HR of 1.6) versus individuals with normal blood pressure. On the other hand, Black race (vs. White) became more prominent for risk for normoglycemic (Figure S8B) individuals. For example, for individuals with FBG 0-100 mg/dL, Black vs. White individuals had a 60% increased risk for diabetes (HR: 1.58, 95% CI: [1.25, 2.00]).

## Conclusions

The clinical utility of the elevated altered glucose tolerance and prediabetes is a hotly debated topic (13). The ADA, in their newest *Standards of Care*, have recommended screening for prediabetes using fasting glucose and A1C as B grade type evidence for normoglycemic individuals. However, their grade was A for recommendation of prevention of cardiovascular disease for individuals who are prediabetics (5); the USPTF have made similar recommendations (14). Our findings indicate that the estimated risk of transition from prediabetes to diabetes is highly dependent on cohort and the chosen definition of prediabetes with inter-cohort range between 2.5 and 15 per 100-person years. Relatedly, the 95% prediction intervals are wide. The observed risk is highest for individuals with FBG between 110-125 mg/dL; however, the inter-cohort range or heterogeneity of the risk is highest for individuals with FBG 110-120 mg/dL (lowest risk for FHS Generation 3 [∼4% annual rate]; highest risk in MESA [∼15%]. Notably, the between cohort range in the incident rates is the lowest in A1C defined prediabetes. Thresholds derived from optimizing diabetes prediction accuracy were similar to the prescribed ADA ones, but accuracy itself varied across cohorts. In summary, there is a need for describing the heterogeneity of risk across diverse cohorts and populations to assess robustness and to communicate the uncertainty (e.g., prediction interval) in the transition rate.

Another point of emphasis includes the difference between the relative risk and transition rate (absolute risk). We observed that overweight and obesity (versus normal weight) and hypertension (versus normal pressure) were associated with higher *relative* rates for diabetes, regardless of individuals being prediabetic or normoglycemic at baseline. The higher and well-known relative risk of obesity for individuals with normoglycemia or prediabetes stands in contrast with the overall transition rate or risk. While relative risks are largest for individuals with obesity across cohorts, regardless of glycemic or prediabetes status, individuals who are obese and are normoglycemic (e.g., HA1C (<=5.7%) or FBG (<=100 mg/dL)) levels transition to diabetes at 1 per 100 person years, among the lowest rates in the distribution we estimated. The transition rates are the lowest for individuals with both lower fasting glucose and HA1C%.

There are a number of implications and strengths in our study. Recently, the USPSTF have acknowledged the need to identify the interindividual or interpopulation sources of the progression to diabetes (17). As we and others highlight, some of the predictors of progression to diabetes may include underlying cardiometabolic risk (e.g., hypertension and overweight/obesity). Second, others have highlighted the progression to diabetes from prediabetes. For example, Koyama et al (6) examine the transition in multiple subgroups of the older population (>65 years of age), documenting annual incidence rates 5-8.2 for individuals with A1C-defined prediabetes. For individuals >65 years of age in the cohorts here, we document a lower incidence rate (overall 1.83, range of 1-4, highest in JHS). On the other hand, Rooney and colleagues (7), for an older subgroup (average age of 75) in the ARIC cohort, document an estimate in line with our own, 1.6 to 2.7. Rooney and colleagues contend that the current prediabetes definition may not be a “robust diagnostic entity” for predicting progression to diabetes in the older population. Our findings support this claim more broadly, finding cohort-level variation may dwarf subgroup effects for diabetes prediction and transition.

Our study highlights the variation of rates based on different thresholds for prediabetes and suggests re-evaluation and/or consideration of the uncertainty in the rate. The thresholds may need to be reevaluated across established complications such as cardiovascular/kidney disease, lower extremity amputation, and other emerging complications, such as cancer(19), with data combined and compared across cohorts. Several systematic reviews and an umbrella review thereof suggest associations of prediabetes with mortality and serious morbidity outcomes (20) (21). However, these associations need to be examined with careful standardization of definitions and with analyses that are not susceptible to selective reporting. Perhaps FBG and/or A1C can be complemented by other risk variables, such as different measures of adiposity (22), participant area-level socioeconomic status or location, that maximize prediction and/or explain the heterogeneity in the trajectory from the existing definition of prediabetes. Estimating cohort-level heterogeneity may help to document uncertainty in the different definitions of prediabetes, a helpful measure to eventually build consensus. These evaluations need to compare effects across different populations and ethnic groups, including those that were not adequately represented in our analyses, e.g. Asian populations (23). Optimizing consensus on definitions is also important in studying intervention effects. Evidence to-date shows no benefit from some widely used interventions, e.g. lifestyle modification, regarding cardiovascular and all-cause mortality (24) and this may be partly due to the difficulty to optimize the definitions in a clinically meaningful way.

The current ADA guidelines deem fasting glucose, A1C, and a 2-hour oral glucose tolerance test as “appropriate” equally in screening for prediabetes and diabetes. Specifically, the criteria include FBG 100-125 mg/dL OR 2-hour OGTT 140-199 mg/dL OR A1C 5.7-6.4%. A1C has been associated with greater incident complications than fasting glucose.(25) Given the large imprecision around all definitions for prediabetes in the transition to diabetes, we suggest studies that embark on examining the (a) consistency – or lack thereof – of the trajectory to diabetes-related complications that move beyond glycemic status (16,26) and/or (b) the appropriate interval for glucose testing after an initial elevated fasting or HA1C test.

## Data Availability

Data are available from the NHLBI BioLINCC repository.

## Author contributions

CJP, WEG, AKM, RV, JPAI designed the study and contributed to drafting of the manuscript.. CJP attained the data and conducted the analysis. CJP takes full responsibility of the work and is the guarantor.

## Supplementary Information

**Table S1.** Sample sizes per cohort of FBG and A1C defined prediabetes subpopulations

**Table S2**. Baseline characteristics per cohort and FBG range.

**Table S3**. Baseline characteristics per cohort and A1C range.

**Table S4.** Subgroup-by-subgroup IR database. Please see the accompanying table. Columns: Weighted IR: incident rate per 100 person-years, subgroup: the subgroup interrogated by BMI, age, sex, or race; IR low: lower 95% CI, IR high: upper 95% CI, pi.lower: lower 95% prediction interval; pi.upper: upper 95% prediction interval

**Table S5**. Optimal cutoffs for fasting glucose and A1C by cohort. Columns: Optimal FBG denotes the threshold picked by the algorithm to maximize accuracy. Sensitivity and Specificity are also described in each column.

**Supplementary Figure 1.** Distributions of baseline fasting glucose A.) Age B.) Race, and C.) Sex; A1C by D.) Age, E.) Race, and F.) Sex

**Supplementary Figure 2.** Rates for incident higher fasting glucose greater than or equal to 126 mg/dL.

**Supplementary Figure 3.** A.) Age and B.) BMI subgroup incidence rate for prediabetes to diabetes per cohort.

**Supplementary Figure 4.** Diabetes Incidence Rates for normoglycemic thresholds for A) Age and B) Body Mass Index ranges.

**Supplementary Figure 5.** Diabetes Incidence Rates for prediabetes and normoglycemic thresholds by sex.

**Supplementary Figure 6.** Prediabetes to diabetes IR by race and cohort. Points are annotated by cohort: ME: MESA; FH: FHS 2; AR: ARIC; JH: JHS

**Supplementary Figure 7.** A.) Hazard Ratio vs. p-value by cohort (row) with prediabetes (FBG 100-125 mg/dL or 110-125 mg/dL, A1C 5.7-6.5%). B.) Hazard Ratio vs. p-value by cohort (row) with normal fasting glucose or glucose control levels (FBG 0-99 mg/dL or 0-109 mg/dL or A1C% 0-5.7). Variables are labeled: Ob: BMI greater than 30 kg/m2 (ref: BMI between 0-25 kg/m2); ow: BMI 25-30 kg/m2 (ref: BMI between 0-25 kg/m2); ht: hypertension; hbp: high blood pressure (ref: normal blood pressure); f: female; Blk: Black race (ref: vs. White); smk: current smoker (ref: never smoker); sage: scaled age; >hs: greater than high school education (versus equivalent to high school education). Framingham 2: Framingham generation 2 cohort; Framingham 3: Framingham generation 3 cohort.

**Supplementary Figure 8.** A.) Meta-analyzed cross-cohort hazard ratio vs. p-value by cohort for individuals with prediabetes (FBG 100-125 mg/dL or 110-125 mg/dL, A1C 5.7-6.5%). B.) Meta-analyzed Hazard ratio vs. p-value by cohort for individuals with normal fasting glucose or glucose control levels (FBG 0-99 mg/dL or 0-109 mg/dL or A1C% 0-5.7). Variables are labeled: Ob: BMI greater than 30 kg/m2 (ref: BMI between 0-25 kg/m2); ow: BMI 25-30 kg/m2 (ref: BMI between 0-25 kg/m2); ht: hypertension; hbp: high blood pressure (ref: normal blood pressure); f: female; Blk: Black race (ref: vs. White); smk: current smoker (ref: never smoker); sage: scaled age; >hs: greater than high school education (versus equivalent to high school education). Framingham 2: Framingham generation 2 cohort; Framingham 3: Framingham generation 3 cohort.

