## Supplementary Document for "Heterogeneity in elevated glucose and A1C as predictors of the prediabetes to diabetes transition: Framingham Heart Study, Multi-Ethnic Study on Atherosclerosis, Jackson Heart Study, and Atherosclerosis Risk In Communities"

### Supplementary Results

#### Incidence rates across subgroups

*Age subgroups*

For the age subgroup, we estimate sex-adjusted rates. Across cohorts (e.g., overall meta-analytic estimate), the FBG range of 110-126 mg/dL had largest overall incidence rates and $I^2$ for both the mid-aged (45-65 years) and older (>65 years of age) individuals (Figure S4A), a meta-analytic estimate of 9% ($I^2$: 99) and 8.1% ($I^2$: 86%) respectively. This is in contrast to individuals with FBG lower than 110 mg/dL, whose overall incidence rate was 1% (Figure S4A) for both the mid-aged and older individuals.

*BMI subgroups*

For the BMI subgroups, the incidence rates we report are adjusted by age and sex. Across cohorts, obese individuals had higher incidence rates across all definitions of prediabetes (Figure S4B) versus individuals who were not obese nor overweight.  The overall/across cohort  incidence rate for FBG 110-125 mg/dL for obese individuals was 9.6 (95% CI: 4.8, 14) per 100-person years ($I^2$: 99%). The incidence rate was approximately half for those under 25 kg/m2 (overall incidence rate of 5 per 100 person-years, $I^2$ 91%) for   the same FBG range (110-125 mg/dL). Furthermore, individuals who were obese but had A1C% in the 5.7-6.5 range had an incidence rate of 3.6 per 100 person-years (95% CI: 1.9-5.3; $I^2$: 95%) (Figure S3B). All the incidence rates for prediabetes ranges, as reflected by their $I^2$, had a high range between cohorts. Take, for example, prediabetes for individuals who were obese and had FBG between 110-126 mg/dL: we observed the maximum incidence rate in the MESA (16.5,  95%CI: 13.9-19.1) and JHS (14.6, 95% CI: 11.2-18.0) population versus for FHS 3 (5, 95%CI: 3-7) (Figure S4B).

The minimum incidence rate of 0.8 per 100 person-years observed for any prediabetes threshold was observed for individuals who had both lower BMI (<25 kg/m2) and had A1C between 5.7-6.5%. For individuals who were obese and had A1C less than 5.7%, the incidence rate was just 1 per 100 person-years ($I^2$: 7%) (Figure S4B). Figure S5 describes incidence rates for age and body mass index for normoglycemic levels. For sex subgroup, we estimated age-adjusted rates and observed a lack of difference between males and females (Figure S6).

*Black and White race/ethnicity subgroups*

The MESA cohorts consisted of participants who were Chinese, White, Hispanic, and Black and the ARIC participants were Black or White (Table S1), in contrast to the FHS and JHS studies, which primarily were composed of Black or White individuals, respectively. We estimated the incidence rate for each cohort and the Black and White race groups (Figure S7). We found that cohort-level differences persisted: black individuals from JHS had higher incidence rate than Black individuals from ARIC; White individuals in MESA had higher incidence rate than White individuals from FHS 2 or 3. Specifically, for FBG 110-125 mg/dL, the incidence rate was 12.5 (95% CI: 9.8, 15) per 100 person-years for JHS (all participants of Black ethnicity) and 15 (95% CI: 12, 18) per 100 person-years for the Black population of MESA. For the Black population in ARIC, however, the incidence rate was only 6 (95% CI: 5.5, 6.8) per 100 person-years (Figure S6). The incidence rate for A1C-based threshold for the Black population in JHS, MESA, and ARIC were 5.2, 1.8, and 1.2 per 100 person-years, respectively. Again, for FBG 110-125 mg/dL, the incidence rate to diabetes for the White subgroup was higher in MESA than in ARIC and FHS: 14 (95% CI: 11, 16.7), 5.3 (95% CI: 4.9, 5.6), and 5% (95% CI: 4.5, 5.5) per 100 person-years, respectively.

**Heterogeneity of data-driven thresholds**

We estimated the heterogeneity of thresholds for fasting glucose and A1C% that are chosen that maximizes the accuracy of prediction of incident diabetes (Table 4). The range of “optimal” thresholds for fasting blood glucose was 95 mg/dL (JHS) to 104 (FHS 2) and for A1C% was 5.6 (ARIC) to 5.8 (JHS and MESA). The $I^2$ for the thresholds was 91% for fasting glucose and 65% for A1C%. The range of accuracies for fasting glucose was 0.73 (ARIC) to 0.88 (MESA).

### Supplementary Tables

**Table S1.** Sample sizes per cohort of FBG and A1C defined prediabetes subpopulations

**Table S2**. Baseline characteristics per cohort and FBG range.

**Table S3**. Baseline characteristics per cohort and A1C range.

**Table S4.** Subgroup-by-subgroup IR database. Please see the accompanying table. Columns: Weighted IR: incident rate per 100 person-years, subgroup: the subgroup interrogated by BMI, age, sex, or race; IR low: lower 95% CI, IR high: upper 95% CI, pi.lower: lower 95% prediction interval; pi.upper: upper 95% prediction interval

**Table S5**. Optimal cutoffs for fasting glucose and A1C by cohort. Columns: Optimal FBG denotes the threshold picked by the algorithm to maximize accuracy. Sensitivity and Specificity are also described in each column.

**Table S1.** Sample sizes per cohort of FBG and A1C defined prediabetes subpopulations

| FBG | ARIC, N = 11,589 | FHS 2, N = 2,171 | FHS 3, N = 2,443 | JHS, N = 1,014 | MESA, N = 5,457 |
|---|---|---|---|---|---|
| 0-109 mg/dL | 10,095 (87%) | 2,098 (97%) | 2,333 (95%) | 992 (98%) | 5,182 (95%) |
| 109-125 mg/dL | 1,494 (13%) | 73 (3.4%) | 110 (4.5%) | 22 (2.2%) | 275 (5.0%) |
| 0-99 mg/dL | 6,508 (56%) | 1,846 (85%) | 1,945 (80%) | 908 (90%) | 4,604 (84%) |
| 100-125 mg/dL | 5,081 (44%) | 325 (15%) | 498 (20%) | 106 (10%) | 853 (16%) |
| Diabetes Diagnosis | 1,801 (16%) | 326 (15%) | 65 (2.7%) | 195 (19%) | 454 (8.3%) |
| A1C | ARIC, N = 4,945 | FHS 2, N = 1,409 | . | JHS, N = 1,083 | MESA, N = 1,971 |
| 0-5.6% | 3,854 (78%) | 1,189 (84%) | | 691 (64%) | 1,484 (75%) |
| 5.7-6.4% | 1,091 (22%) | 220 (16%) | | 392 (36%) | 487 (25%) |
| Diabetes Diagnosis | 842 (17%) | 62 (4.4%) | | 191 (18%) | 155 (7.9%) |

**Table S2**. Baseline characteristics per cohort and FBG range.

| | ARIC | | FHS 2 | | FHS 3 | | JHS | | MESA | |
|---|---|---|---|---|---|---|---|---|---|---|
| **Glucose Range (mg/dL)** | **0-99** | **100-125** | **0-99** | **100-125** | **0-99** | **100-125** | **0-99** | **100-125** | **0-99** | **100-125** |
| **Fasting Glucose mg/dL (IQR)** | 93 (89, 96) | 105 (102, 111) | 88 (84, 93) | 104 (101, 109) | 90 (86, 94) | 104 (101, 108) | 87 (82, 91) | 104 (101, 108) | 86 (81, 91) | 106 (102, 112) |
| **SEX** | | | | | | | | | | |
| Male | 2,470 (38%) | 2,721 (54%) | 800 (43%) | 203 (62%) | 759 (39%) | 365 (73%) | 344 (38%) | 47 (44%) | 2,076 (45%) | 488 (57%) |
| Female | 4,038 (62%) | 2,360 (46%) | 1,046 (57%) | 122 (38%) | 1,186 (61%) | 133 (27%) | 564 (62%) | 59 (56%) | 2,528 (55%) | 365 (43%) |
| **Age** | | | | | | | | | | |
| 18-44 | 24 (0.4%) | 9 (0.2%) | 878 (48%) | 80 (25%) | 1,370 (70%) | 252 (51%) | 359 (40%) | 18 (17%) | 6 (0.1%) | 0 (0%) |
| 45-64 | 6,454 (99%) | 5,030 (99%) | 926 (50%) | 227 (70%) | 564 (29%) | 239 (48%) | 480 (53%) | 71 (67%) | 2,773 (60%) | 406 (48%) |
| 64- | 30 (0.5%) | 42 (0.8%) | 42 (2.3%) | 18 (5.5%) | 11 (0.6%) | 7 (1.4%) | 69 (7.6%) | 17 (16%) | 1,825 (40%) | 447 (52%) |
| **Education** | | | | | | | | | | |
| High School | 2,203 (34%) | 1,667 (33%) | 559 (30%) | 114 (35%) | 228 (12%) | 100 (20%) | 135 (15%) | 22 (21%) | 808 (18%) | 165 (19%) |
| < High School | 1,193 (18%) | 1,082 (21%) | 56 (3.0%) | 16 (4.9%) | 13 (0.7%) | 9 (1.8%) | 59 (6.5%) | 13 (12%) | 643 (14%) | 191 (22%) |
| > High School | 3,112 | 2,332 | 1,231 | 195 | 1,704 | 389 | 714 | 71 | 3,153 | 497 |

| | | | | | | | | | | |
|---|---|---|---|---|---|---|---|---|---|---|
| | (48%) | (46%) | (67%) | (60%) | (88%) | (78%) | (79%) | (67%) | (68%) | (58%) |
| **Race** | | | | | | | | | | |
| White | 5,188 (80%) | 4,040 (80%) | 1,846 (100%) | 325 (100%) | 1,869 (96%) | 470 (94%) | 0 (0%) | 0 (0%) | 2,047 (44%) | 264 (31%) |
| Chinese | 0 (0%) | 0 (0%) | 0 (0%) | 0 (0%) | 29 (1.5%) | 8 (1.6%) | 0 (0%) | 0 (0%) | 504 (11%) | 124 (15%) |
| Black | 1,320 (20%) | 1,041 (20%) | 0 (0%) | 0 (0%) | 15 (0.8%) | 7 (1.4%) | 908 (100%) | 106 (100%) | 1,141 (25%) | 253 (30%) |
| Hispanic | 0 (0%) | 0 (0%) | 0 (0%) | 0 (0%) | 32 (1.6%) | 13 (2.6%) | 0 (0%) | 0 (0%) | 912 (20%) | 212 (25%) |
| **Smoking** | | | | | | | | | | |
| No | 4,882 (75%) | 3,906 (77%) | 1,356 (73%) | 263 (81%) | 1,707 (88%) | 430 (86%) | 810 (89%) | 92 (87%) | 4,012 (87%) | 749 (88%) |
| Current | 1,626 (25%) | 1,175 (23%) | 490 (27%) | 62 (19%) | 238 (12%) | 68 (14%) | 98 (11%) | 14 (13%) | 592 (13%) | 104 (12%) |
| **Body Mass Index** | | | | | | | | | | |
| Median (IQR) | 25.6 (23.1, 28.7) | 27.5 (24.9, 30.9) | 24.8 (22.3, 27.6) | 27.4 (25.2, 30.7) | 25.1 (22.4, 28.4) | 28.7 (25.8, 32.4) | 30 (26, 35) | 34 (31, 39) | 26.9 (24.1, 30.3) | 29.0 (26.4, 32.9) |
| 0-24 kg/m2 | 2,846 (44%) | 1,309 (26%) | 959 (52%) | 76 (23%) | 949 (49%) | 90 (18%) | 137 (15%) | 2 (1.9%) | 1,527 (33%) | 131 (15%) |
| 25-29 kg/m2 | 2,474 (38%) | 2,222 (44%) | 662 (36%) | 153 (47%) | 675 (35%) | 220 (44%) | 301 (33%) | 24 (23%) | 1,833 (40%) | 365 (43%) |
| >=30 kg/m2 | 1,188 (18%) | 1,550 (31%) | 225 (12%) | 96 (30%) | 321 (17%) | 188 (38%) | 470 (52%) | 80 (75%) | 1,244 (27%) | 357 (42%) |
| **Blood Pressure** | | | | | | | | | | |
| Normal | 4,484 (69%) | 3,038 (60%) | 1,035 (56%) | 102 (31%) | 1,379 (71%) | 226 (45%) | 535 (59%) | 47 (44%) | 2,774 (60%) | 395 (46%) |
| High BP (SBP >= 130 or DBP >-=80) | 911 (14%) | 861 (17%) | 387 (21%) | 75 (23%) | 305 (16%) | 105 (21%) | 176 (19%) | 23 (22%) | 725 (16%) | 160 (19%) |
| Hypertension (SBP >= 140 or SBP >= 85) | 1,113 (17%) | 1,182 (23%) | 424 (23%) | 148 (46%) | 261 (13%) | 167 (34%) | 197 (22%) | 36 (34%) | 1,105 (24%) | 298 (35%) |

**Table S3**. Baseline characteristics per cohort and A1C range.

| | **ARIC** | | **FHS 2** | | **JHS** | | **MESA** | |
|---|---|---|---|---|---|---|---|---|
| **A1C Range (%)** | **0-5.6** | **5.7-6.4** | **0-5.6** | **5.7-6.4** | **0-5.6** | **5.7-6.4** | **0-5.6** | **5.7-6.4** |
| **A1C%** | 5.30 (5.10, | 5.80 (5.70, | 5.07 (4.79, | 5.89 (5.79, 6.07) | 5.30 (5.10, | 5.90 (5.80, | 5.30 (5.10, | 5.80 (5.70, 6.00) |

| | | | | | | | | |
|---|---|---|---|---|---|---|---|---|
| | 5.40) | 6.00) | 5.33) | | 5.50) | 6.10) | 5.50) | |
| **SEX** | | | | | | | | |
| Male | 1,605 (42%) | 491 (45%) | 508 (43%) | 98 (45%) | 272 (39%) | 155 (40%) | 815 (55%) | 269 (55%) |
| Female | 2,249 (58%) | 600 (55%) | 681 (57%) | 122 (55%) | 419 (61%) | 237 (60%) | 669 (45%) | 218 (45%) |
| **Age** | | | | | | | | |
| 18-44 | 0 (0%) | 0 (0%) | 226 (19%) | 21 (9.5%) | 290 (42%) | 96 (24%) | 0 (0%) | 0 (0%) |
| 45-64 | 3,701 (96%) | 1,030 (94%) | 830 (70%) | 143 (65%) | 353 (51%) | 252 (64%) | 926 (62%) | 247 (51%) |
| 64- | 153 (4.0%) | 61 (5.6%) | 133 (11%) | 56 (25%) | 48 (6.9%) | 44 (11%) | 558 (38%) | 240 (49%) |
| **Education** | | | | | | | | |
| High School | 1,318 (34%) | 373 (34%) | 358 (30%) | 73 (33%) | 105 (15%) | 69 (18%) | 223 (15%) | 110 (23%) |
| < High School | 378 (9.8%) | 211 (19%) | 40 (3.4%) | 17 (7.7%) | 43 (6.2%) | 32 (8.2%) | 153 (10%) | 68 (14%) |
| > High School | 2,158 (56%) | 507 (46%) | 791 (67%) | 130 (59%) | 543 (79%) | 291 (74%) | 1,108 (75%) | 309 (63%) |
| **Race** | | | | | | | | |
| White | 3,398 (88%) | 698 (64%) | 1,189 (100%) | 220 (100%) | 0 (0%) | 0 (0%) | 836 (56%) | 153 (31%) |
| Chinese | 0 (0%) | 0 (0%) | 0 (0%) | 0 (0%) | 0 (0%) | 0 (0%) | 73 (4.9%) | 26 (5.3%) |
| Black | 456 (12%) | 393 (36%) | 0 (0%) | 0 (0%) | 691 (100%) | 392 (100%) | 326 (22%) | 201 (41%) |
| Hispanic | 0 (0%) | 0 (0%) | 0 (0%) | 0 (0%) | 0 (0%) | 0 (0%) | 249 (17%) | 107 (22%) |
| **Smoking** | | | | | | | | |
| No | 3,310 (86%) | 862 (79%) | 1,001 (84%) | 173 (79%) | 619 (90%) | 338 (86%) | 1,178 (79%) | 383 (79%) |
| Current | 544 (14%) | 229 (21%) | 188 (16%) | 47 (21%) | 72 (10%) | 54 (14%) | 306 (21%) | 104 (21%) |
| **Body Mass Index** | | | | | | | | |
| 0-24 kg/m2 | 1,495 (39%) | 236 (22%) | 472 (40%) | 56 (25%) | 117 (17%) | 27 (6.9%) | 454 (31%) | 86 (18%) |
| 25-29 kg/m2 | 1,607 (42%) | 468 (43%) | 482 (41%) | 90 (41%) | 249 (36%) | 120 (31%) | 627 (42%) | 175 (36%) |
| >=30 kg/m2 | 752 (20%) | 387 (35%) | 235 (20%) | 74 (34%) | 325 (47%) | 245 (62%) | 403 (27%) | 226 (46%) |
| **Blood Pressure** | | | | | | | | |
| Normal | 2,893 (75%) | 698 (64%) | 726 (61%) | 107 (49%) | 416 (60%) | 199 (51%) | 1,003 (68%) | 279 (57%) |

| | | | | | | | | |
|---|---|---|---|---|---|---|---|---|
| High BP (SBP >= 130 or DBP >-=80) | 487 (13%) | 187 (17%) | 211 (18%) | 41 (19%) | 129 (19%) | 92 (23%) | 215 (14%) | 95 (20%) |
| Hypertension (SBP >= 140 or SBP >= 85) | 474 (12%) | 206 (19%) | 252 (21%) | 72 (33%) | 146 (21%) | 101 (26%) | 266 (18%) | 113 (23%) |

**Table S4.** Subgroup-by-subgroup IR database. Please see the accompanying table. Columns: Weighted IR: incident rate per 100 person-years, subgroup: the subgroup interrogated by BMI, age, sex, or race; IR low: lower 95% CI, IR high: upper 95% CI, pi.lower: lower 95% prediction interval; pi.upper: upper 95% prediction interval

**Table S5**. Optimal cutoffs for fasting glucose and A1C by cohort. Columns: Optimal FBG denotes the threshold picked by the algorithm to maximize accuracy. Sensitivity and Specificity are also described in each column.

| **Cohort** | **Optimal FBG Threshold (CI)** | **Optimal A1C Threshold (CI)** | **Accuracy FBG** | **Sensitivity FBG** | **Specificity FBG** | **Accuracy A1C** | **Sensitivity A1C** | **Specificity A1C** |
|---|---|---|---|---|---|---|---|---|
| **FHS 2** | 104 (102.3, 105.7) | 5.53 (5.14, 5.92) | 0.84 | 0.8 | 0.84 | 0.76 | 0.56 | 0.77 |
| **FHS 3** | 101 (99.1, 102.9) | . | 0.84 | 0.75 | 0.85 | . | . | . |
| **MESA** | 103 (100.0, 106.0) | 5.8 (5.62, 5.98) | 0.88 | 0.69 | 0.89 | 0.83 | 0.54 | 0.85 |
| **JHS** | 95 (92.1, 97.9) | 5.8 (5.65, 5.95) | 0.71 | 0.65 | 0.71 | 0.73 | 0.75 | 0.73 |
| **ARIC** | 103.9 (102.1, 105.7) | 5.6 (5.55, 5.65) | 0.73 | 0.74 | 0.73 | 0.7 | 0.53 | 0.74 |
| **Overall (CI) I2** | 101.5 (98.29, 104.67); 91% | 5.7 (5.56,5.82); 65% | | | | | | |

### Supplementary Figures

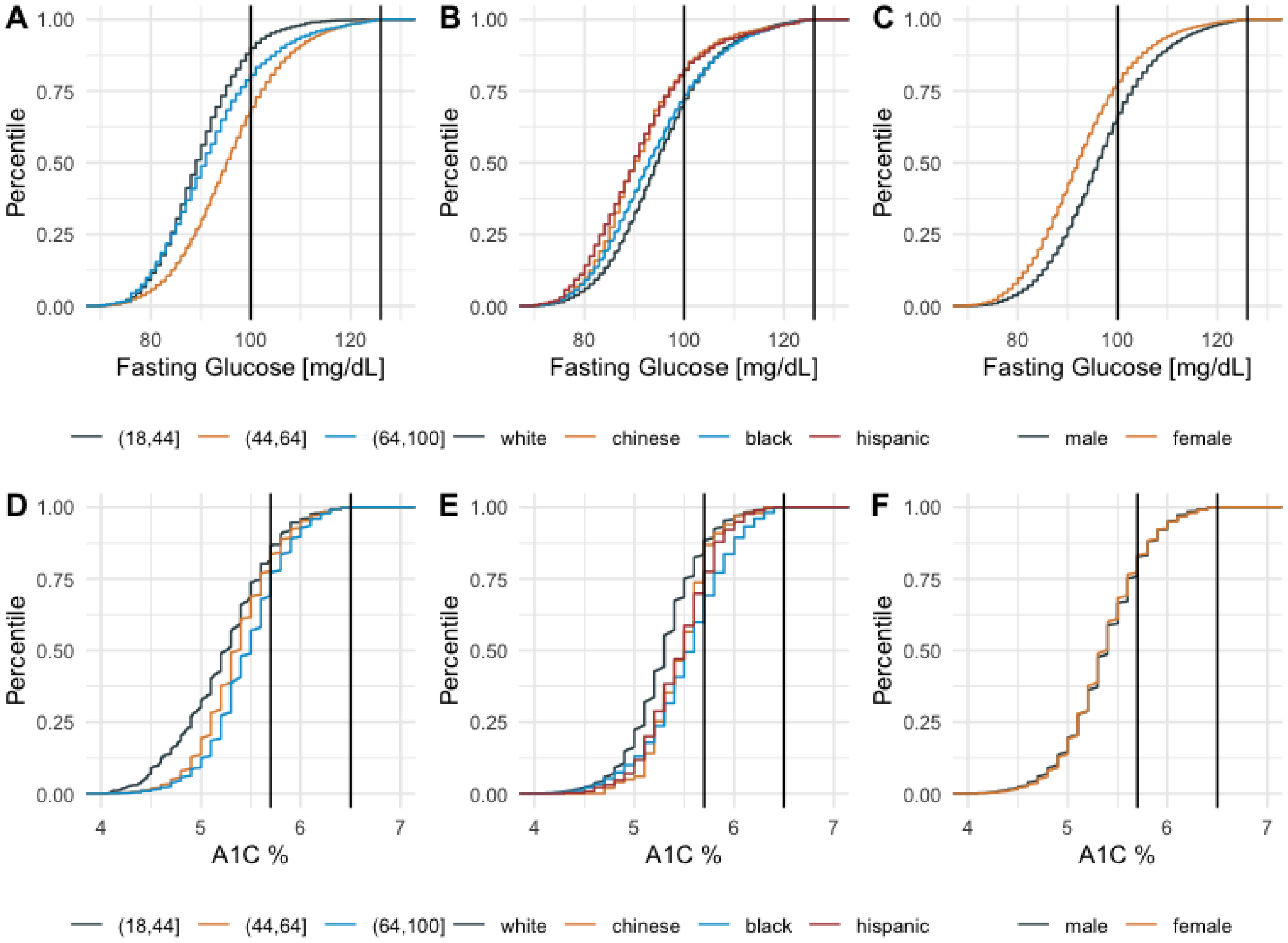


**Supplementary Figure 1**. Distributions of baseline fasting glucose A.) Age B.) Race, and C.) Sex and A1C by D.) Age, E.) Race, and F.) Sex

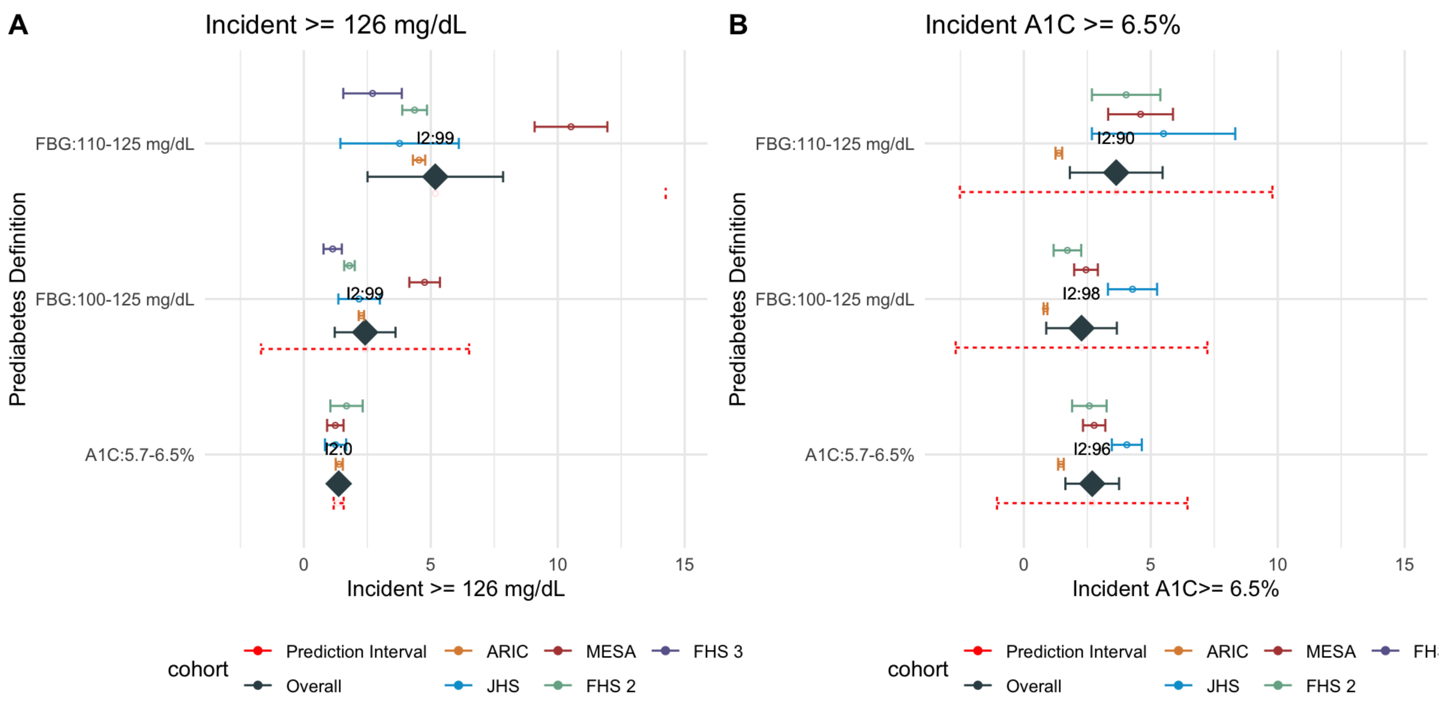


**Supplementary Figure 2.** Rates for incident higher fasting glucose greater than or equal to 126 mg/dL.

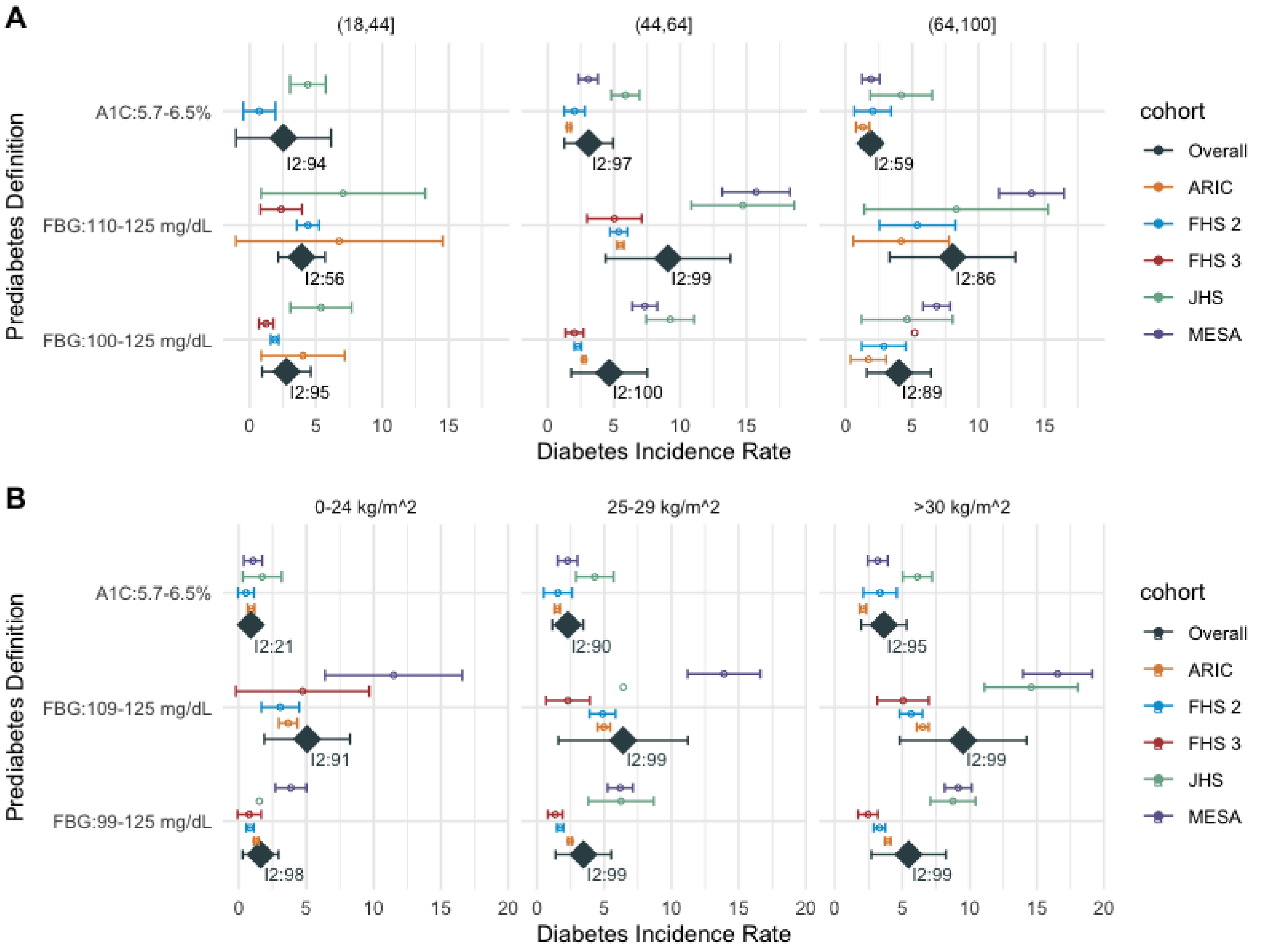


**Supplementary Figure 3**. A.) Age and B.) BMI subgroup incidence rate for prediabetes to diabetes per cohort.

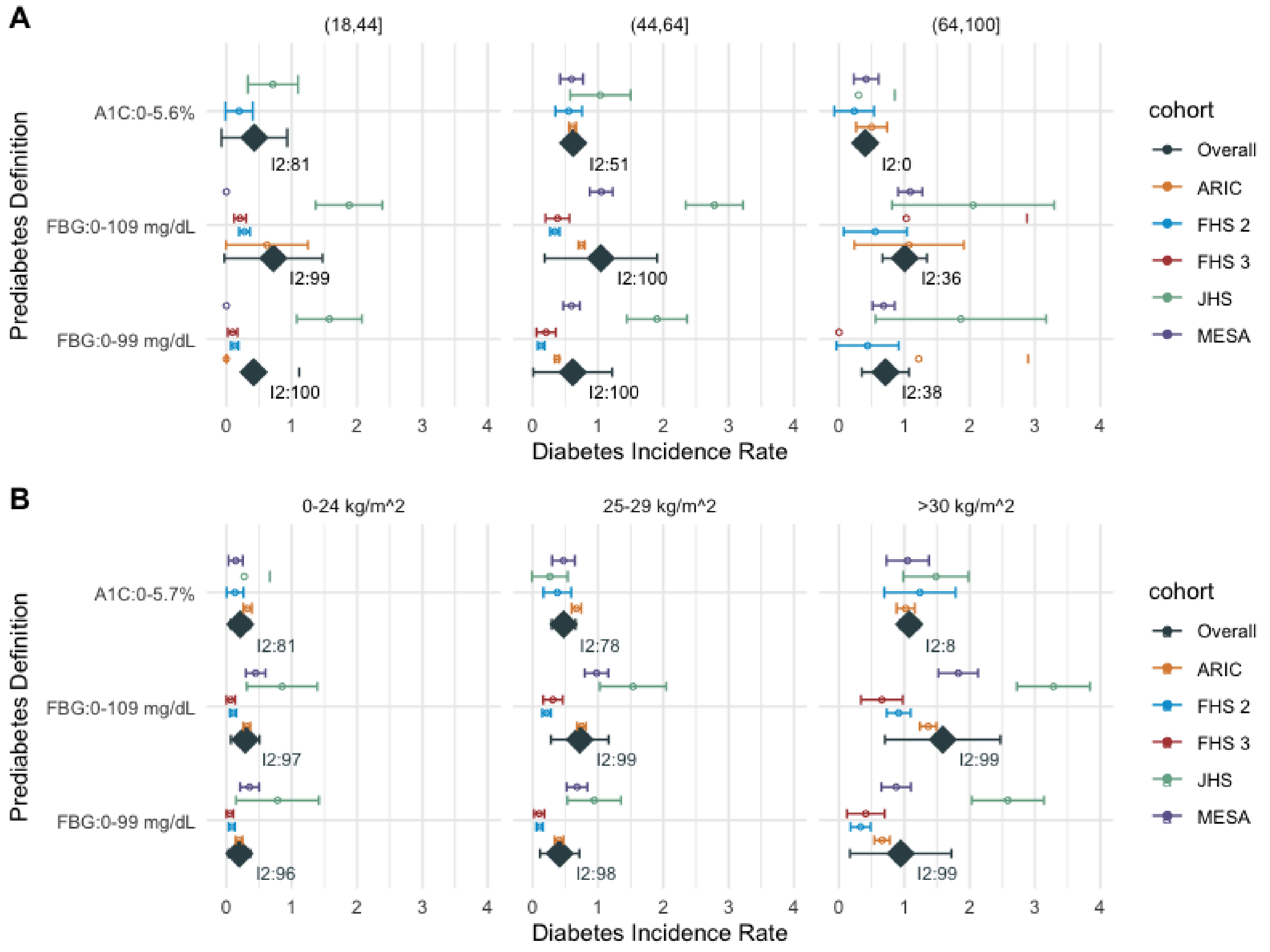


**Supplementary Figure 4.** Diabetes Incidence Rates for normoglycemic thresholds for A) Age and B) Body Mass Index ranges.

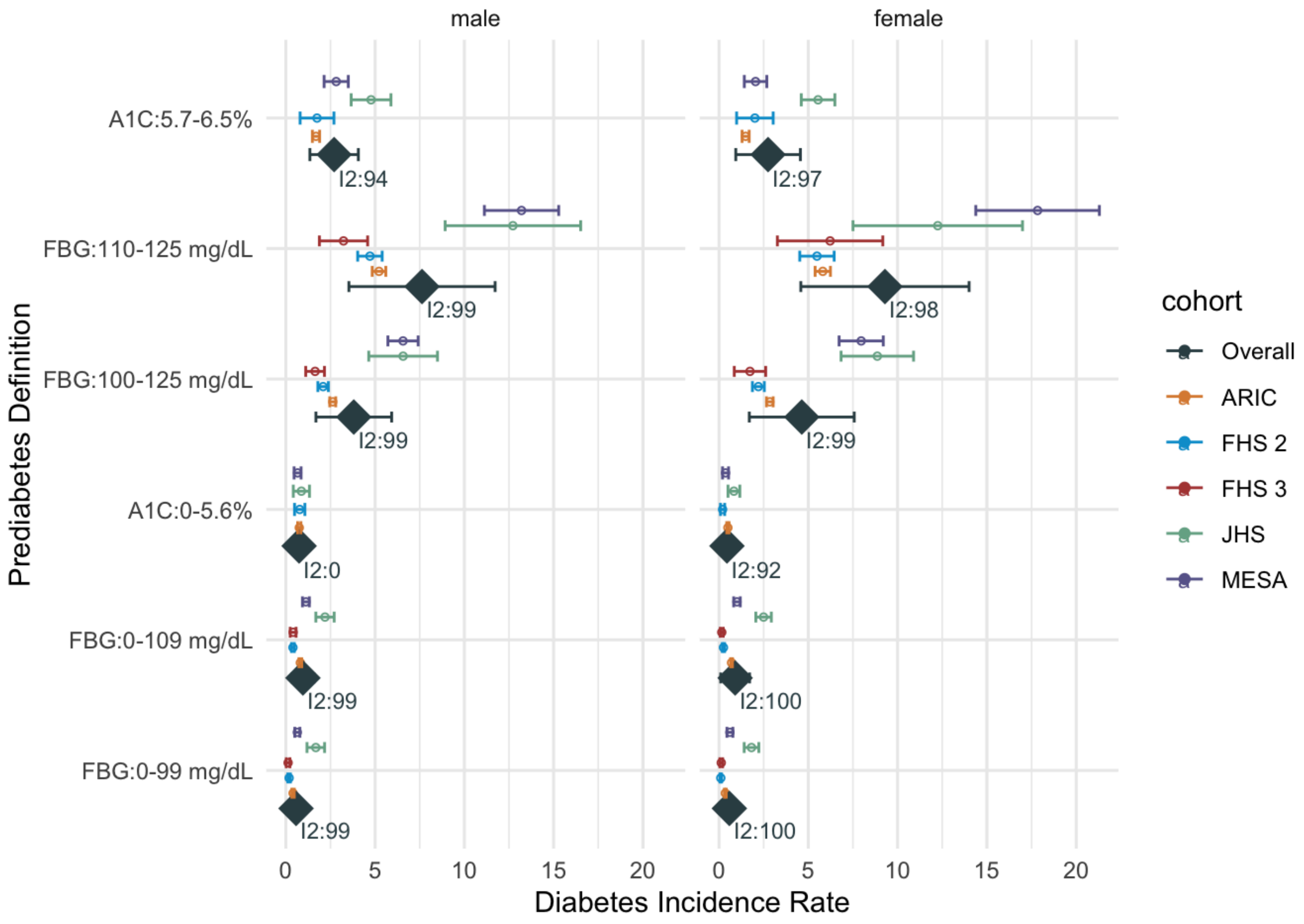


**Supplementary Figure 5**. Diabetes Incidence Rates for prediabetes and normoglycemic thresholds by sex.

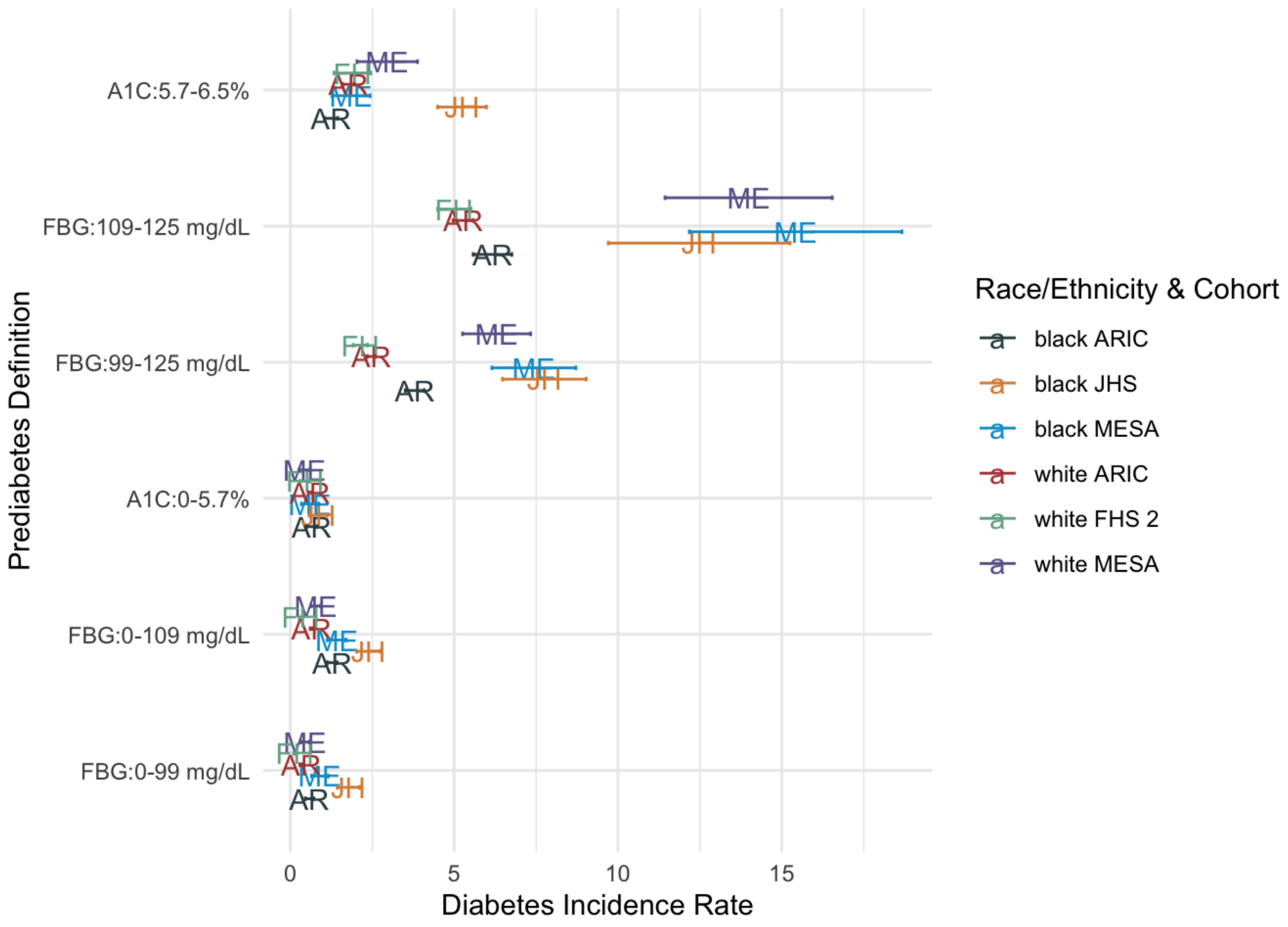


**Supplementary Figure 6**. Prediabetes to diabetes IR by race and cohort. Points are annotated by cohort: ME: MESA; FH: FHS 2; AR: ARIC; JH: JHS

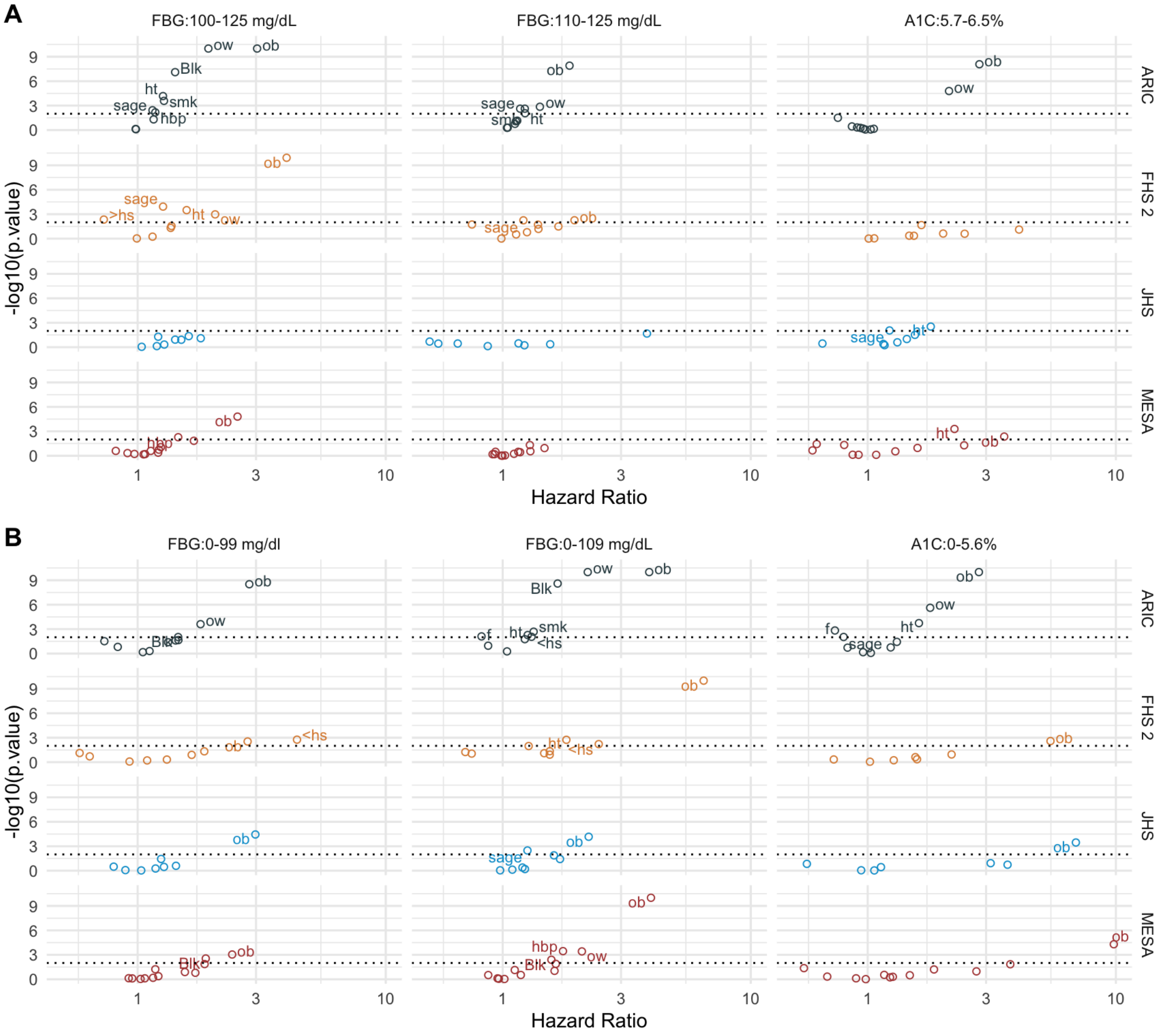


**Supplementary Figure 7.** A.) Hazard Ratio vs. p-value by cohort (row) with prediabetes (FBG 100-125 mg/dL or 110-125 mg/dL, A1C 5.7-6.5%). B.) Hazard Ratio vs. p-value by cohort (row) with normal fasting glucose or glucose control levels (FBG 0-99 mg/dL or 0-109 mg/dL or A1C% 0-5.7). Variables are labeled: Ob: BMI greater than 30 kg/m2 (ref: BMI between 0-25 kg/m2); ow: BMI 25-30 kg/m2 (ref: BMI between 0-25 kg/m2); ht: hypertension; hbp: high blood pressure (ref: normal blood pressure); f: female; Blk: Black race (ref: vs. White); smk: current smoker (ref: never smoker); sage: scaled age; >hs: greater than high school education (versus equivalent to high school education). Framingham 2: Framingham generation 2 cohort; Framingham 3: Framingham generation 3 cohort.

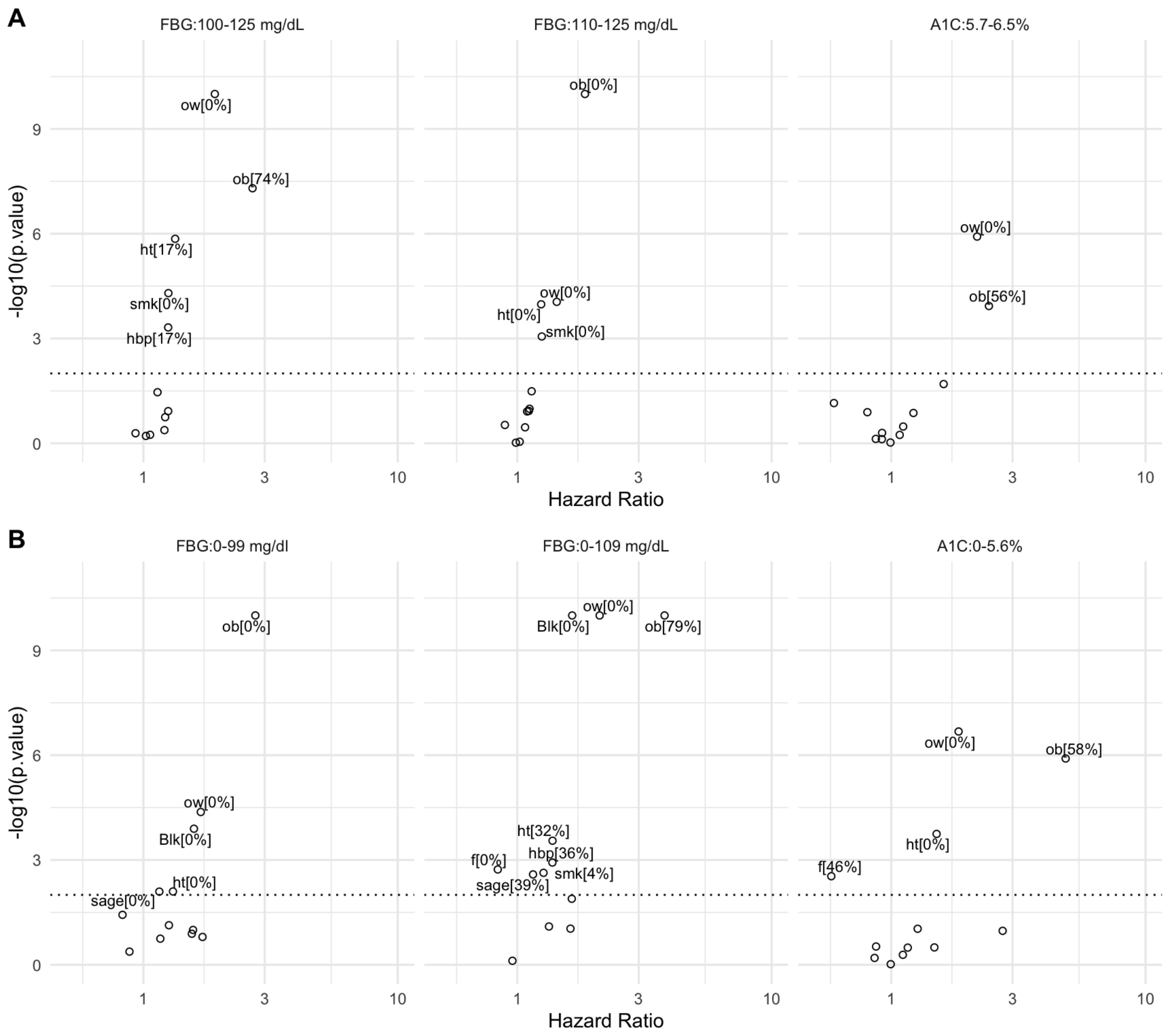


**Supplementary Figure 8.** A.) Meta-analyzed cross-cohort hazard ratio vs. p-value by cohort for individuals with prediabetes (FBG 100-125 mg/dL or 110-125 mg/dL, A1C 5.7-6.5%). B.) Meta-analyzed Hazard ratio vs. p-value by cohort for individuals with normal fasting glucose or glucose control levels (FBG 0-99 mg/dL or 0-109 mg/dL or A1C% 0-5.7). Variables are labeled: Ob: BMI greater than 30 kg/m2 (ref: BMI between 0-25 kg/m2); ow: BMI 25-30 kg/m2 (ref: BMI between 0-25 kg/m2); ht: hypertension; hbp: high blood pressure (ref: normal blood pressure); f: female; Blk: Black race (ref: vs. White); smk: current smoker (ref: never smoker); sage: scaled age; >hs: greater than high school education (versus equivalent to high school education). Framingham 2: Framingham generation 2 cohort; Framingham 3: Framingham generation 3 cohort.
